# Cardiac rehabilitation for outpatients aged over 80 years with cardiovascular diseases

**DOI:** 10.1101/2025.02.20.25322642

**Authors:** Ryoko Someya, Yasushi Matsuzawa, Yoshitaka Shimizu, Hidefumi Nakahashi, Masaaki Konishi, Eiichi Akiyama, Yohei Hanajima, Hisaya Kondo, Tomohiro Yoshii, Ryosuke Sato, Kazuko Hayashi, Kozo Okada, Teruyasu Sugano, Kiyoshi Hibi

**Affiliations:** Department of Rehabilitation, Yokohama City University Medical Center, Yokohama, Japan; Department of Cardiovascular Medicine, Graduate School of Medical Sciences, Kumamoto University, Kumamoto, Japan; Division of Cardiology, Yokohama City University Medical Center, Yokohama, Japan; Department of Medical Science and Cardiorenal Medicine, Yokohama City University Graduate School of Medicine, Yokohama, Japan; Kawaguchi Cardiovascular and Respiratory Hospital, Saitama, Japan

**Author notes:** **Corresponding author:** Yasushi Matsuzawa, MD, PhD, Department of Cardiovascular Medicine, Graduate School of Medical Sciences, Kumamoto, University. 1-1-1 Honjo, Chuo-ku, Kumamoto, 860-8556, Japan.

**Keywords:** Older adults, Cardiovascular disease, Cardiac Rehabilitation, Cardiopulmonary exercise test, Frailty, Vascular endothelial function

## Abstract

**Background:** In 2023, Japan had the highest proportion of the elderly in the world, with one in 10 individuals aged ≥80 years. Consequently, the number of patients with cardiovascular diseases is increasing. Older patients often have comorbidities such as frailty, sarcopenia, and cognitive decline that leads to a decreased quality of life (QOL). We analyzed the effects of outpatient cardiac rehabilitation (OCR) in elderly patients with cardiovascular disease.

**Methods:** The comprehensive OCR had been provided by a multidisciplinary team. We analyzed data from 49 patients with cardiovascular diseases, aged ≥80 years, who received cardiopulmonary exercise (CPX) tests based OCR. Frailty, physical function, QOL, exercise tolerance, and vascular endothelial function were assessed before and after OCR.

**Results:** All 49 patients had completed the OCR program. The mean patients’ age was 84.1 ± 3.6 years and 32.7% were male. The proportion of frailty and pre-frailty participants significantly decreased from 92% to 67% following OCR. The QOL (KCCQ: 76.8 ± 18.4 vs. 81.4 ± 20.4; P = 0.0196), exercise tolerance (peak VO₂: 14.1 ± 4.0 vs. 15.4 ± 3.9 mL/min/kg; P = 0.0017), and vascular endothelial function (Ln-RHI: 0.48 ± 0.39 vs. 0.57 ± 0.3; P = 0.027) significantly improved after OCR.

**Conclusions:** The comprehensive OCR with CPX-based exercise therapy and the multidisciplinary approach significantly improved frailty, the QOL, physical function, exercise tolerance, and vascular endothelial function in patients with cardiovascular diseases aged ≥80 years.

## Introduction

Populations are rapidly aging worldwide, especially in Japan, where the proportion of the older population is the highest in the world, with 1 in 10 citizens being ≥80 years.^1^ According to the Japanese Registry of All Cardiac and Vascular Diseases (JROAD), patients’ population with heart disease is also aging.^2,3^

Comorbidities in older patients, such as frailty, sarcopenia, and dementia, contribute to decreased exercise tolerance and worsening quality of life (QOL). The prevalence of frailty increases with age and significantly influences the prognosis of patients with cardiovascular diseases.^4,5^ In fact, 56.1% of older patients with heart failure in Japan are physically frail.^5^ In addition to physical decline, frailty is also associated with malnutrition, mental and cognitive impairment, and other comorbidities, necessitating cardiac rehabilitation (CR) with a comprehensive multidisciplinary approach.^6^

The benefits of comprehensive CR for patients with cardiovascular diseases are well established. Consequently, it is a Class I recommendation in the United States of America, Europe, and Japan.^7,8,9^ CR improves exercise tolerance, reduces cardiovascular mortality, decreases readmissions, enhances QOL, and furthermore improves vascular endothelial function,^9,10,11,12,13^ with these benefits also observed in patients with cardiovascular diseases, aged ≥70years, regarding exercise tolerance, QOL, and frailty. ^14,15,16^ However, studies focusing on older patients, particularly those ≥80 years, are limited, with few studies cardiopulmonary exercise tests (CPX) or following guidelines principles.^9^

Therefore, this study aimed to determine the effects of outpatient CR(OCR) on frailty, QOL, exercise tolerance, and vascular endothelial function among patients aged ≥80 years with cardiovascular diseases.

## Methods

This retrospective, observational study analyzed data from patients (n = 67; age: ≥80 years) with cardiovascular diseases who were admitted or attended to Yokohama City University Medical Center between December 2020 and December 2022. All participated in CPX before and after OCR.

The Yokohama City University Ethics Committee approved this study (Approval ID: F240500014). Informed consent was obtained under the approval of the ethics committee and innominate data were analyzed. All parameters below were assessed at baseline and after participating in the OCR program.

### Outpatient CR

The frequency of 1-h sessions was once or twice per week for 3–4 months. The intensity was determined based on the anaerobic threshold (AT) determined with the CPX. When the AT could not be evaluated due to oscillation, a perceived exertion level of 11–13 on the Borg scale was used instead. The exercise regimen comprised 20 min of stretching and resistance training using body weight or weighted resistance, followed by 30 min of aerobic exercise on a stationary bicycle ergometer. Doctors, physical therapists and nurses educated the patients about heart disease, mainly focusing on fostering self-monitoring habits, interpreting results, and providing lifestyle guidance and exercise instruction. Nurses provided individual counseling sessions for patients with depressive tendencies or other reasons. Dietitians tailored individual nutritional guidance according to dietary intake and exercise levels.

### Physical function and body composition

Grip strength was measured using a dynamometer (Takei Scientific Instruments Co., Ltd., Niigata, Japan). The knee extension strength was assessed using a handheld dynamometer (Anima Corporation, New York, NY, USA). The maximum values of two measurements were recorded. Knee extension strength was calculated as a ratio (%) of body weight (%BW). Walking speed was assessed by measuring comfortable walking speed twice, and the faster speed was recorded. The Short Physical Performance Battery (SPPB) was applied and a 6-min walk distance (6MWD) was measured on a 40 m walking course. Skeletal muscle mass (SMI) measured using an InBody770 body composition analyzer (InBody, Tokyo, Japan)

### Frailty and sarcopenia

Frailty was assessed using the revised Japanese version of the Cardiovascular Health Study (J-CHS) criteria.^17^ Individuals meeting ≥3 criteria were classified as frail, while those meeting 1–2 criteria were classified as pre-frail. Sarcopenia was evaluated baseline and after participating in the OCR program based on SMI, grip strength, and walking speed according to the criteria of the Asian Working Group for Sarcopenia 2019 (AWGS2019) .^18^

### Cognitive function, depression, ADL, QOL

Cognitive function was assessed using the Mini-Mental State Examination-Japanese (MMSE-J), ^19^ and mental function was evaluated using the Patient Health Questionnaire-9 (PHQ-9). ^20^ Activities of daily living were assessed using the Barthel Index (BI). QOL was evaluated using the Kansas City Cardiomyopathy Questionnaire (KCCQ). ^21^

### Cardiopulmonary exercise tests

Cardiopulmonary exercise tolerance in CPX was analyzed using the breath-by-breath method with an expiratory gas analyzer (Minato Medical Science Co., Ltd., Osaka, JAPAN) and a Strength Ergo cycle ergometer. The ramp protocol comprised rest for 3 min, a 3 min warm-up (0–10 watts), and a ramp increase of 10 W/min. Peak VO₂ was defined as oxygen consumption at the peak exercise workload, and the AT was determined using the V-slope method.

### Endothelial function test

Vascular endothelial function was assessed by measuring the reactive hyperemia index (RHI), which detects the arterial dilation response to brachial artery occlusion as a fingertip volume pulse wave. This was automatically measured using an Endo-PAT2000 device (Zoll Itamar Ltd., Atlanta, GA, USA) baseline and after participation in the OCR program. The RHI was transformed using the natural logarithm (Ln-RHI) and utilized for analysis.

### Statistical analysis

Results are presented as n (%), means ± standard deviations (SD), or as medians with interquartile ranges (IQR), depending on the nature and distribution of the variables. Parameters were compared at baseline and after participation in the OCR program using paired t-tests and Chi-squared tests. All data were statistically analyzed using JMP Pro 17. Values with P < 0.05 were considered statistically significant.

## Results

Among 67 patients included in the study, 62 participated after hospital discharge, while five were outpatients. Among the 67 patients, 14 dropped out and four had missing data, resulting in our analysis comprising 49 patients (average age, 84.1 ± 3.6 years; male, 32.7%; mean body mass index, 22.9 ± 3.8 kg/m²) (**Figure 1**). During the same period, 486 (28%) of the 1,738 patients admitted to our cardiovascular center were aged ≥80 years, of whom 258 received CR during hospitalization and 62 continued with OCR. Diagnoses included myocardial infarction (12.2%), angina pectoris (4.1%), heart failure (22.5%), transcatheter aortic valve implantation (TAVI) (57.1%), and lower extremity arterial disease (LEAD) (4.1%) (**Table 1**).

**Figure 1.**
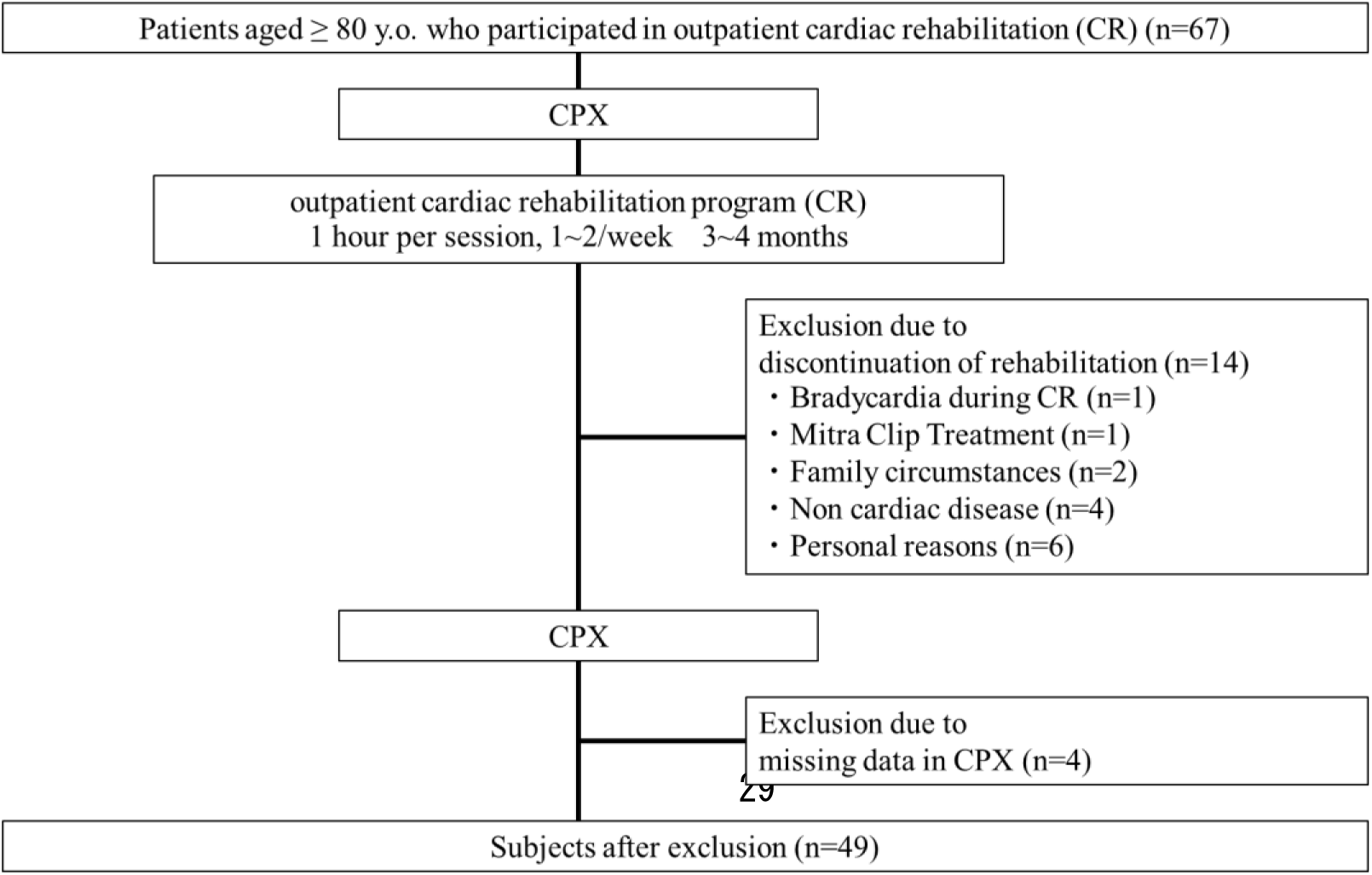
Flow chart of patient enrollment and evaluation. CPX, cardiopulmonary exercise tests; OCR, outpatient cardiovascular rehabilitation.

**Table1.**
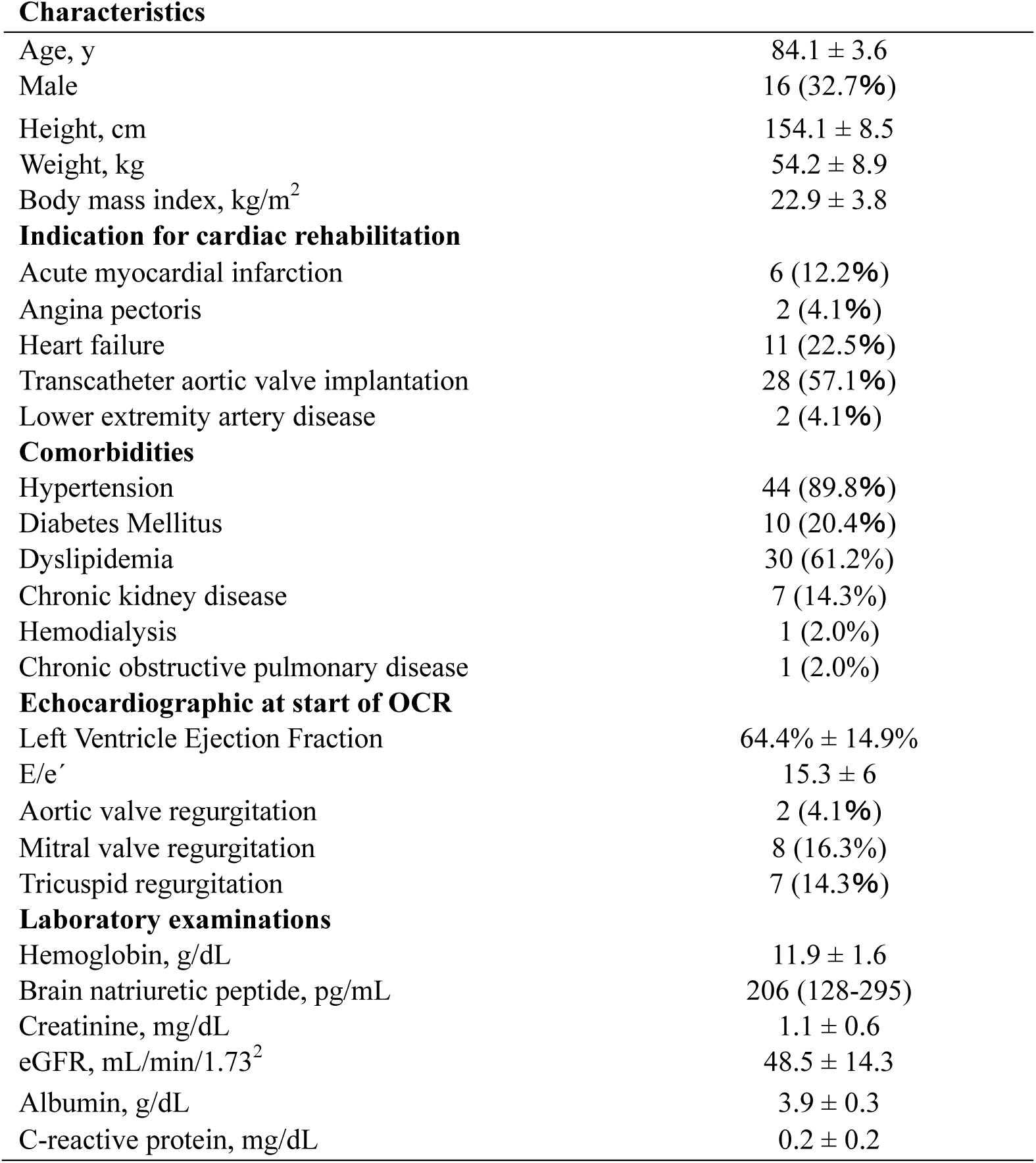
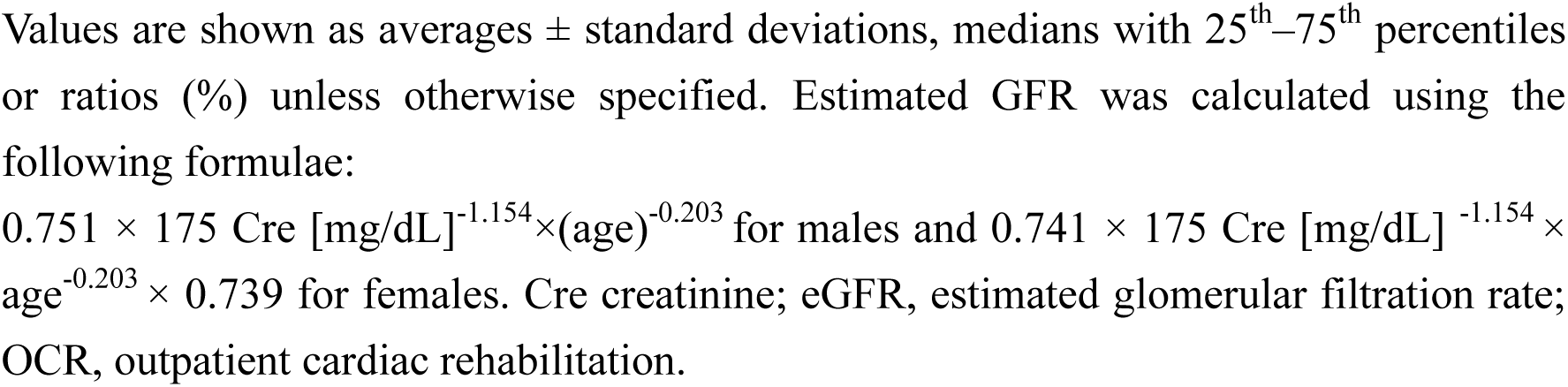
Characteristics of Study Population (n = 49).

| **Characteristics** |  |
| --- | --- |
| Age, y | $84.1 \pm 3.6$ |
| Male | 16 (32.7%) |
| Height, cm | $154.1 \pm 8.5$ |
| Weight, kg | $54.2 \pm 8.9$ |
| Body mass index, $kg/m^2$ | $22.9 \pm 3.8$ |
| **Indication for cardiac rehabilitation** |  |
| Acute myocardial infarction | 6 (12.2%) |
| Angina pectoris | 2 (4.1%) |
| Heart failure | 11 (22.5%) |
| Transcatheter aortic valve implantation | 28 (57.1%) |
| Lower extremity artery disease | 2 (4.1%) |
| **Comorbidities** |  |
| Hypertension | 44 (89.8%) |
| Diabetes Mellitus | 10 (20.4%) |
| Dyslipidemia | 30 (61.2%) |
| Chronic kidney disease | 7 (14.3%) |
| Hemodialysis | 1 (2.0%) |
| Chronic obstructive pulmonary disease | 1 (2.0%) |
| **Echocardiographic at start of OCR** |  |
| Left Ventricle Ejection Fraction | $64.4\% \pm 14.9\%$ |
| E/e' | $15.3 \pm 6$ |
| Aortic valve regurgitation | 2 (4.1%) |
| Mitral valve regurgitation | 8 (16.3%) |
| Tricuspid regurgitation | 7 (14.3%) |
| **Laboratory examinations** |  |
| Hemoglobin, g/dL | $11.9 \pm 1.6$ |
| Brain natriuretic peptide, pg/mL | 206 (128-295) |
| Creatinine, mg/dL | $1.1 \pm 0.6$ |
| eGFR, $mL/min/1.73^2$ | $48.5 \pm 14.3$ |
| Albumin, g/dL | $3.9 \pm 0.3$ |
| C-reactive protein, mg/dL | $0.2 \pm 0.2$ |

Values are shown as averages ± standard deviations, medians with 25<sup>th</sup>–75<sup>th</sup> percentiles or ratios (%) unless otherwise specified. Estimated GFR was calculated using the following formulae:
$0.751 \times 175 \text{ Cre [mg/dL]}^{-1.154} \times (\text{age})^{-0.203}$ for males and $0.741 \times 175 \text{ Cre [mg/dL]}^{-1.154} \times \text{age}^{-0.203} \times 0.739$ for females. Cre creatinine; eGFR, estimated glomerular filtration rate; OCR, outpatient cardiac rehabilitation.

### Outpatient CR

A total of 32 (65%) patients participated in OCR sessions twice a week. Aerobic exercise at the AT level was achieved by 23 (46.9%) patients. Lower limb fatigue or decreased muscular endurance prevented the others from achieving the AT, thus their exercise intensity was adjusted to a Borg scale rating of 11–13. The reasons for discontinuation of OCR in the 14 excluded patients were bradycardia during outpatient OCR (n = 1), Transcatheter edge to edge repair treatment during OCR (n = 1), family circumstances (n = 2), non-cardiac diseases (n = 4), and personal reasons (n = 6). Consequently, data from 49 patients were analyzed.

### Findings at baseline and after OCR

**Table 2** and **Figures 2** and **3** show the results of comparisons between baseline and after OCR. Physical function significantly improved after OCR. Knee extension strength increased from 20.0 ± 7.8 to 21.9 ± 6.9 kg force (kgf) (P < 0.01) and SPPB scores improved from 10.1 ± 2.7 to 10.7 ± 2.0 (P < 0.01). Walking speed increased from 0.88 ± 0.3 to 0.95 ± 0.3 m/s (P < 0.01) and 6MWD improved from 360 ± 123 to 403 ± 103 m (P < 0.001). The proportion of frailty and pre-frailty patients, assessed using the J-CHS, decreased from 91.8% to 67.3% (P =0.0015); **Figure 2**. The MMSE-J score also significantly improved.

**Figure 2.**
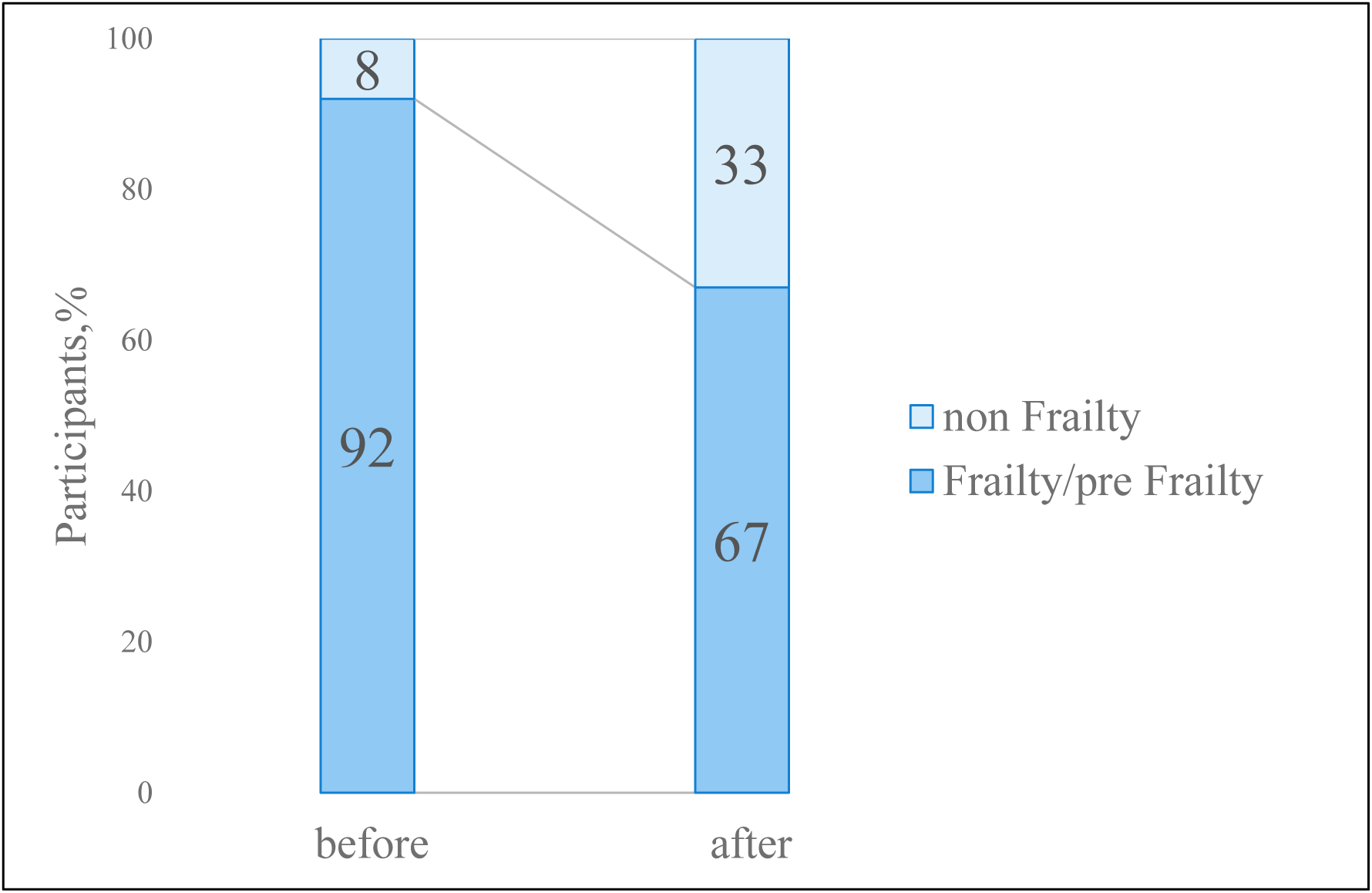
Changes in frailty/pre-frailty between baseline and after OCR. (91.8% vs. 67.3%, P = 0.0015). OCR, outpatient cardiovascular rehabilitation.

**Figure 3.**
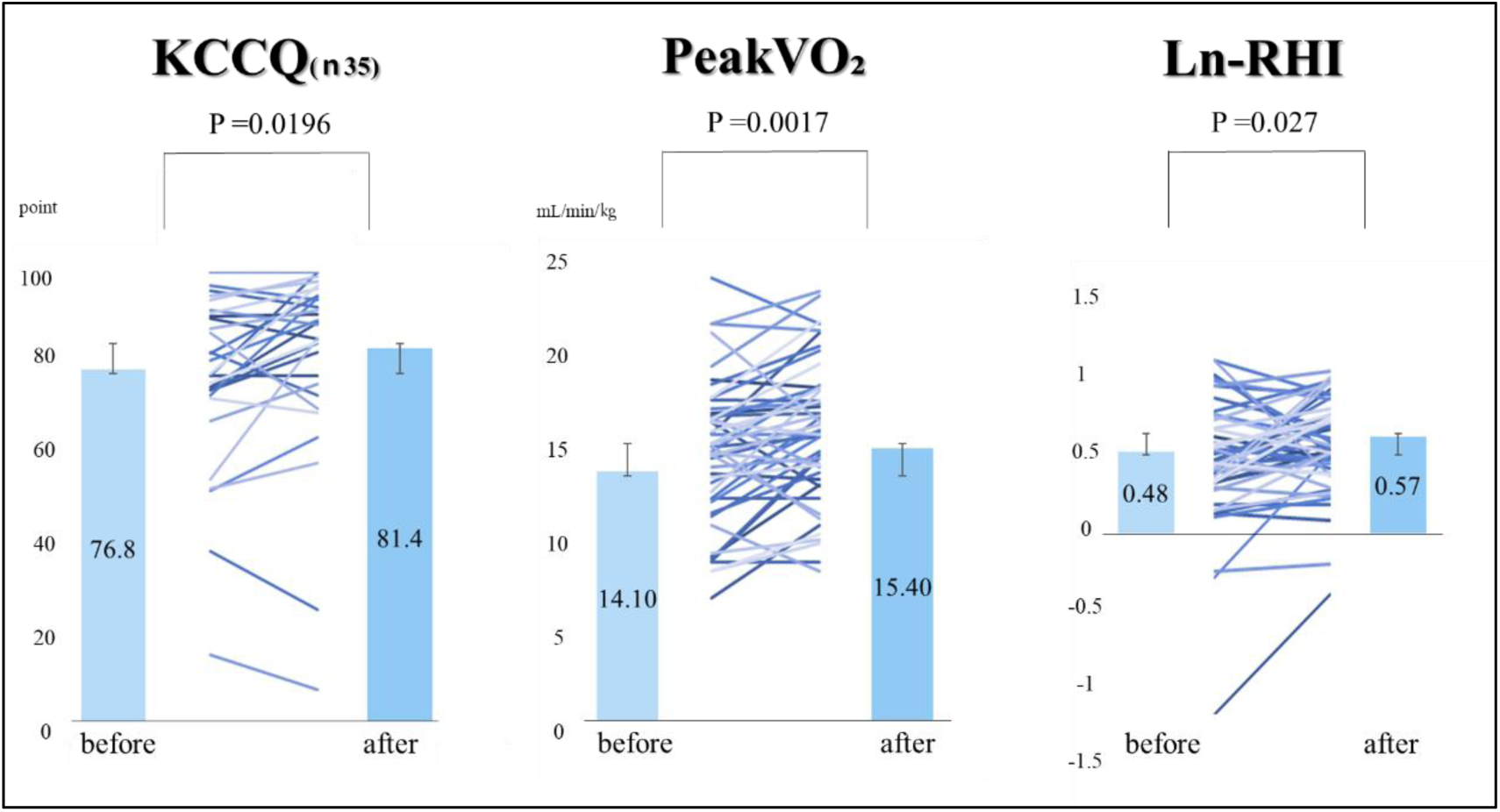
Changes in KCCQ, peak KCCQ, LnRHI, at baseline and after OCR. KCCQ (76.8 ± 18.4 *vs*. 81.4 ± 20.4; P = 0.0196), peak VO₂ (14.1 ± 4.0 *vs*. 15.4 ± 3.9 mL/min/ ㎏; P = 0.0017), LnRHI (0.48 ± 0.39 *vs*. 0.57 ± 0.3; P = 0.027). KCCQ, Kansas City Cardiomyopathy Questionnaire; LnRHI, natural logarithmic transformation of the reactive hyperemia index; OCR, outpatient cardiovascular rehabilitation; VO₂, volume of oxygen per unit of time.

**Table 2.**
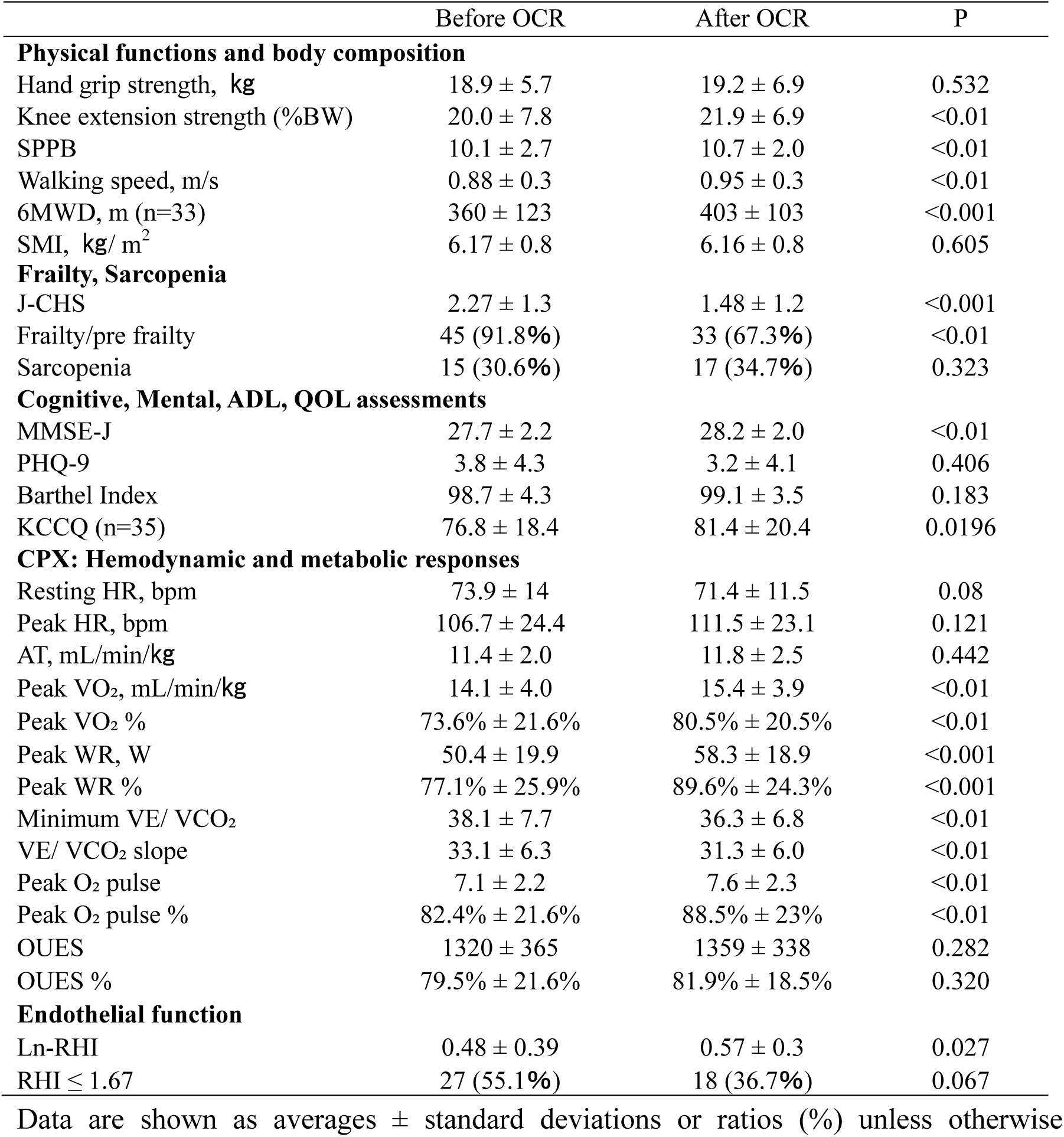
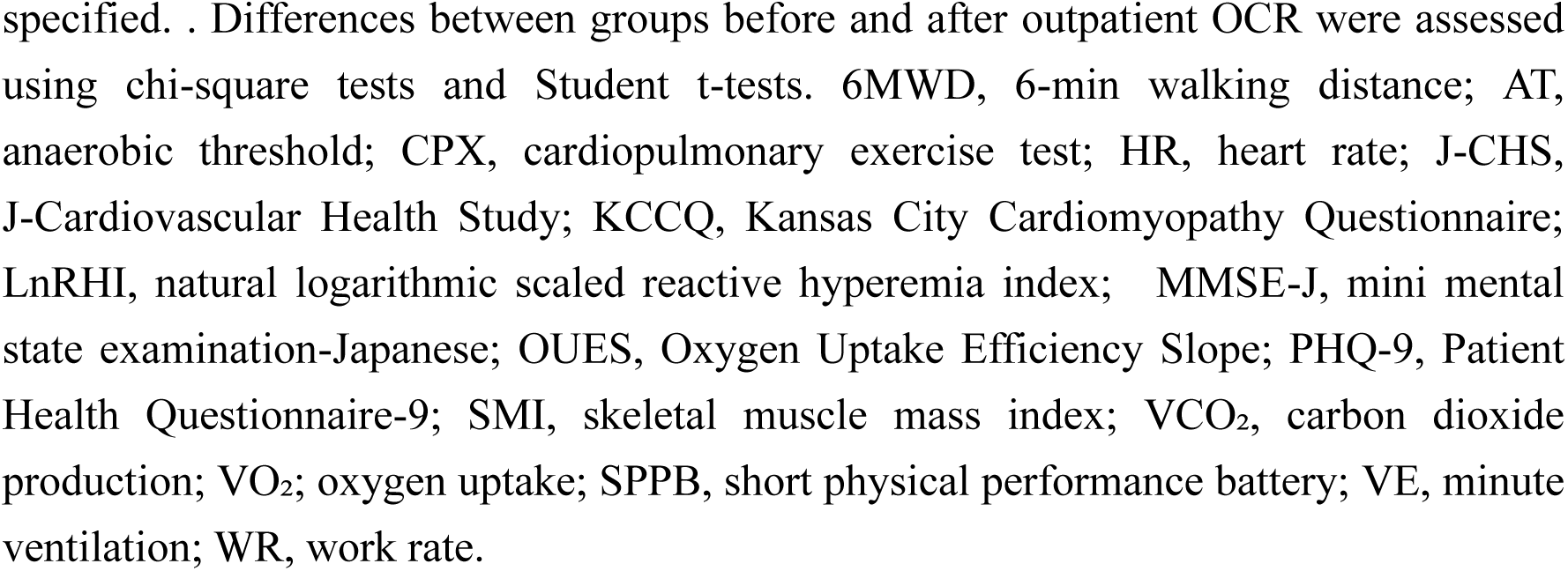
Measured parameters.

|  | Before OCR | After OCR | P |
| --- | --- | --- | --- |
| **Physical functions and body composition** |  |  |  |
| Hand grip strength, kg | 18.9 ± 5.7 | 19.2 ± 6.9 | 0.532 |
| Knee extension strength (%BW) | 20.0 ± 7.8 | 21.9 ± 6.9 | <0.01 |
| SPPB | 10.1 ± 2.7 | 10.7 ± 2.0 | <0.01 |
| Walking speed, m/s | 0.88 ± 0.3 | 0.95 ± 0.3 | <0.01 |
| 6MWD, m (n=33) | 360 ± 123 | 403 ± 103 | <0.001 |
| SMI, kg/ m<sup>2</sup> | 6.17 ± 0.8 | 6.16 ± 0.8 | 0.605 |
| **Frailty, Sarcopenia** |  |  |  |
| J-CHS | 2.27 ± 1.3 | 1.48 ± 1.2 | <0.001 |
| Frailty/pre frailty | 45 (91.8%) | 33 (67.3%) | <0.01 |
| Sarcopenia | 15 (30.6%) | 17 (34.7%) | 0.323 |
| **Cognitive, Mental, ADL, QOL assessments** |  |  |  |
| MMSE-J | 27.7 ± 2.2 | 28.2 ± 2.0 | <0.01 |
| PHQ-9 | 3.8 ± 4.3 | 3.2 ± 4.1 | 0.406 |
| Barthel Index | 98.7 ± 4.3 | 99.1 ± 3.5 | 0.183 |
| KCCQ (n=35) | 76.8 ± 18.4 | 81.4 ± 20.4 | 0.0196 |
| **CPX: Hemodynamic and metabolic responses** |  |  |  |
| Resting HR, bpm | 73.9 ± 14 | 71.4 ± 11.5 | 0.08 |
| Peak HR, bpm | 106.7 ± 24.4 | 111.5 ± 23.1 | 0.121 |
| AT, mL/min/kg | 11.4 ± 2.0 | 11.8 ± 2.5 | 0.442 |
| Peak VO<sub>2</sub>, mL/min/kg | 14.1 ± 4.0 | 15.4 ± 3.9 | <0.01 |
| Peak VO<sub>2</sub> % | 73.6% ± 21.6% | 80.5% ± 20.5% | <0.01 |
| Peak WR, W | 50.4 ± 19.9 | 58.3 ± 18.9 | <0.001 |
| Peak WR % | 77.1% ± 25.9% | 89.6% ± 24.3% | <0.001 |
| Minimum VE/ VCO<sub>2</sub> | 38.1 ± 7.7 | 36.3 ± 6.8 | <0.01 |
| VE/ VCO<sub>2</sub> slope | 33.1 ± 6.3 | 31.3 ± 6.0 | <0.01 |
| Peak O<sub>2</sub> pulse | 7.1 ± 2.2 | 7.6 ± 2.3 | <0.01 |
| Peak O<sub>2</sub> pulse % | 82.4% ± 21.6% | 88.5% ± 23% | <0.01 |
| OUES | 1320 ± 365 | 1359 ± 338 | 0.282 |
| OUES % | 79.5% ± 21.6% | 81.9% ± 18.5% | 0.320 |
| **Endothelial function** |  |  |  |
| Ln-RHI | 0.48 ± 0.39 | 0.57 ± 0.3 | 0.027 |
| RHI ≤ 1.67 | 27 (55.1%) | 18 (36.7%) | 0.067 |

Data are shown as averages ± standard deviations or ratios (%) unless otherwise
specified. . Differences between groups before and after outpatient OCR were assessed using chi-square tests and Student t-tests. 6MWD, 6-min walking distance; AT, anaerobic threshold; CPX, cardiopulmonary exercise test; HR, heart rate; J-CHS, J-Cardiovascular Health Study; KCCQ, Kansas City Cardiomyopathy Questionnaire; LnRHI, natural logarithmic scaled reactive hyperemia index; MMSE-J, mini mental state examination-Japanese; OUES, Oxygen Uptake Efficiency Slope; PHQ-9, Patient Health Questionnaire-9; SMI, skeletal muscle mass index; VCO<sub>2</sub>, carbon dioxide production; VO<sub>2</sub>; oxygen uptake; SPPB, short physical performance battery; VE, minute ventilation; WR, work rate.

The CPX data revealed significant improved peak VO₂, peak work rate (WR), peak O₂ pulse. Ventilation efficiency and indices of heart failure severity such as minimum VE/VCO₂ and VE/VCO₂ slope were also improved.

Endothelial function, Ln-RHI, significantly improved, and the frequency of endothelial dysfunction defined as RHI ≤1.67 tended to decrease. **Figure 3** shows the results of KCCQ (76.8 ± 18.4 vs. 81.4 ± 20.4, P =0.0196), peak VO₂ (14.1 ± 4.0 vs. 15.4 ± 3.9 mL/min/kg, P = 0.0017), and Ln-RHI (0.48 ± 0.39 vs. 0.57 ± 0.3, P =0.027). QOL, exercise tolerance, and endothelial function significantly improved after OCR.

### Adverse events related to OCR

Serious complications or sudden conditional changes did not develop during the CPX tests. One patient developed bradycardia during the OCR and the session was interrupted. In another case, heart failure worsened during treatment, requiring medication adjustments; regardless, the patient was able to continue cardiac rehabilitation after being discharged from the hospital.

## Discussion

Our findings revealed that OCR significantly reduced the prevalence of frailty and pre-frailty, and significantly improved QOL, exercise tolerance, and endothelial function in patients with cardiovascular diseases aged ≥80 years.

We found that the incidence of frailty and pre-frailty significantly decreased from 92% to 67% after OCR. Similarly, The REHAB-HF trial, which involved patients with heart failure with an average age of 75 ± 6 years, found a 97% prevalence of frailty and pre-frailty.^16^ Although the patients in the present study were older than those in the REHAB-HF trial, frailty obviously improved following OCR. The J-CHS criteria, based on a proposed definition of physical frailty,^22^ assesses frailty as weight loss, muscle weakness, exhaustion, slow walking speed, and low levels of physical activity. Furthermore, frailty encompasses cognitive, nutritional, and psychosocial factors, all of which require targeted interventions.^6^ The OCR program provided in this study adhered to set guidelines, incorporating CPX-based aerobic exercise and resistance training tailored to each individual.^9^ Additionally, we provided patient education, counseling, and nutritional guidance through a multidisciplinary approach. This comprehensive strategy allowed us to address not only the physical components of frailty, but also other domains, leading to a multifaceted improvement in frailty. In older patients with cardiac disease and frailty, comprehensive CR, which includes multidisciplinary interventions, appears to be more effective than exercise therapy alone.

In this study, QOL was significantly improved in 71% of the participants following OCR. Japanese guidelines also recommend CR for older patients with coronary artery disease and heart failure to improve their QOL.^9^ In patients with heart failure, combining aerobic exercise with resistance training improves peak VO₂, muscle strength, and the QOL.^23^ A KCCQ score improvement of over 5 points has been associated with reduced mortality in heart failure hospitalization.^21^ However, a recent study indicated that even improvements below 5 points may have clinical significance.^24^ In this study, the overall increase in the KCCQ score was 4.6 points, a statistically significant improvement, suggesting that CR contributed to the desired improvement in QOL for older patients. However, some patients showed a significant decline in the KCCQ score compared to the baseline, highlighting the need for further investigation into background factors and appropriate interventions.

According to the CPX results, parameters including the peak VO₂ and ventilatory efficiency improved. In patients with cardiovascular disease, peak VO₂ is associated with prognosis, with an improvement of 3.5 mL/kg/min (1 Met) corresponding to a 12% reduction in mortality.^25^ In the HF-ACTION trial for patients with chronic heart failure, a 6% improvement in peak VO₂ was identified as the threshold for reducing mortality.^26^ Although the present study did not achieve an improvement of 1 Met in peak VO₂, it did show a 9.2% improvement, suggesting that despite the older age of the participants, CR can enhance exercise tolerance and potentially improve prognosis. Minimum VE/VCO₂ and VE/VCO₂ slope are known prognostic indicators for HFrEF,^27^ and in patients aged 80 and over post-TAVI, the VE/VCO₂ slope is associated with heart failure re-hospitalization.^28^ In this study, these indices also showed significant improvement, suggesting the potential for CR to alleviate dyspnea, the primary symptom of heart failure, improve prognosis, and reduce the likelihood of rehospitalization. Exercise prescription based on CPX results is considered safe and is recommended by guidelines.^9,29^ However, the application of CPX-based CR in older patients with frailty is often limited due to comorbidities and decreased balance capabilities. Despite the inclusion of several frail patients in this study, no major adverse events occurred during CPX or OCR, indicating that OCR was performed safely. This suggests that even among the older population, conducting evaluations and exercise prescriptions based on CPX with adequate risk management is highly beneficial.

We found that OCR significantly improved endothelial function, as assessed by the RHI, in older patients. Endothelial function, evaluated by flow-mediated vasodilation or RHI, predicts cardiovascular events and is associated with prognosis.^30^ Specifically, RHI is considered a prognostic factor for patients with heart failure,^31,32^ and resistance training improves endothelial function in patients with cardiac disease.^33^ Moreover, patients with chronic heart failure who regularly exercise have improved endothelial function that correlates with peak VO₂.^34^ We found that regular resistance training and aerobic exercise during OCR significantly improved skeletal muscle function, as evidenced by increased peak VO₂ and peak WR, along with enhanced endothelial function.

In this study only 12.8% of hospitalized patients aged ≥80 years participated in the OCR sessions, among whom 65% twice weekly. Approximately ∼20% of patients in need are referred to rehabilitation in the USA, among whom only approximately ∼30% participate in OCR.^35^ The rates of patients in Japan with acute coronary syndromes or heart failure who participate in OCR were reportedly 10%^36^ and 7%^37^, respectively. Although the participation rate and frequency were high in the present study, it highlights the need for further improvement in participation rates. The barriers to OCR participation are low referral rates, inadequate physician awareness, gender disparity, physical discomfort, socioeconomic and psychological factors, and distance.^38^ Our patients were aged ≥80 years, and factors such as decreased adherence, time constraints due to treatment of comorbidities, and difficulty with frequent outpatient visits, particularly for those living alone, were significant factors that limited participation and led to dropout. Future efforts should focus on improving adherence and establishing support systems for outpatient visits, while also considering the feasibility of home-based CR programs.

### Limitations

This study was a single-center, retrospective analysis of a small sample of patients who could participate in OCR and thus lacked a control group. Subgroup analyses for each disease could not be determined due to the size of the sample. Despite the older age of our patients, they were relatively high-functioning and were capable of outpatient visits. Therefore, we did not include patients with lower ADLs who would be unable to participate in OCR. Future studies should include patients with extreme mobility limitations and explore appropriate interventions. Although the KCCQ revealed significant overall improvements, it was primarily designed for patients with stage C or worse heart failure with reduced ejection fraction (HFrEF). Here, the KCCQ was applied to patients with heart failure stages ranging from A to C, which might not be appropriate. Furthermore, we did not follow up the patients over periods or analyze clinical events following the completion of the OCR sessions, thus emphasizing the need for further investigation in these areas. Our patients were evaluated from the late recovery period when they were stable after inpatient treatment or TAVI. It has been reported that improvements in QOL at 1 and 12 months,^39^ along with immediate and delayed improvements in vascular endothelial function.^40^ Since approximately 60% of the patients were post-TAVI, we cannot rule out the possibility that these factors may have influenced the changes observed after OCR.

## Conclusion

We concluded that comprehensive OCR that includes exercise therapy based on CPX and multidisciplinary approaches significantly contributes to improving frailty, QOL, physical function, exercise tolerance, and endothelial function in patients aged ≥80 years with cardiovascular diseases. Including OCR in treatment regimens in addition to devices and pharmacological approaches, is a fundamental strategy for significantly enhancing the prognoses and QOL of older patients with cardiovascular diseases.

## Data Availability

The data produced and analyzed in this study are available from the corresponding author upon reasonable request.

## Acknowledgement

None.

